# Persisting low skin prick test prevalence among children receiving periodic anthelminthic treatment after recurrent soil-transmitted helminth infections: a one-year observation

**DOI:** 10.1101/2022.10.10.22280898

**Authors:** Inke Nadia D. Lubis, Masitah Nasution, Rizky Keumala Ansari Nasution, Aridamuriany D. Lubis, Gema Nazri Yanni, Yunnie Trisnawati, Ferryan Sofyan

## Abstract

**Background:** The immune system in human develops when they are exposed to severe helminth infections. Chronic soil-transmitted helminth (STH) infection can modulate and suppress allergic reactions particularly by changing the responses from the immune effector. This study observed how STH infections are correlated with allergic reactions as determined by skin prick test (SPT).

**Methodology:** Fourty-five primary school children with recurrent STH infections (determined by at least two positive results in 4-monthly Kato Katz examination over a year period) who received periodic treatment from Mandailing Natal, were classified as case, and 45 primary school students with no history of STH infections from Medan were classified as control in this study. Positive SPT occurred among 27% and 89% of children in the case and control groups (OR 22, 95% CI 7.032-68.827), respectively. No history of helminthiasis, history of allergy, history of parent’s allergies, and history of sibling’s allergies were associated with increased risks of positive SPT.

**Conclusion:** Our findings supported the hygiene hypothesis, where decreased exposure to infectious disease pathogens and decreased diversity of microbial exposures in the environment increases the prevalence of allergies, and anthelminthic treatment show limited effect in reversing the protective effect of STH infections to allergies.

## Introduction

Allergies have become more common over the world, where the prevalence has dramatically increased from 30% to 40% in both developed and developing countries [1]. Allergies among the urban populations are assumed to be related to the westernized lifestyle. On the other hand, the rural population in the developing countries are more vulnerable to the microorganisms in the environment that protect them from allergies and atopic condition [2,3]. The hygiene hypothesis has been proposed to explain the relationship between the decreased exposure to common childhood infectious diseases and the decreased diversity of microbial exposures in the environment, such as geohelminth (*Ascaris lumbricoides, Trichuris trichiura*, and hookworms) infections and the increasing prevalence of allergies [1,3].

The host immune provides similar responses to either the environmental allergens or parasite antigens by secreting high IgE levels, tissue eosinophilia and mastocytosis, mucus hypersecretion, and T cells that preferentially secrete Th2 cytokines (IL-4, IL-5, dan IL-13) [4,5,6,7]. Chronic soil-transmitted helminth (STH) infections can modulate and suppress allergic reactions, particularly by changing the responses from the immune effector and secreting immunomodulator cytokines that stimulate the protective dominant T-helper cell type-2 (Th2), recruit the immune system cells, such as eosinophils, basophils, mast cells, and macrophages, and differential production of immunoglobulin, such as IgG1, IgG4, and especially IgE, which are the same elements as the allergic responses [4,5]. Children whose allergic inflammatory responses are controlled by peripheral tolerance to cross-reactive antigens may keep suppressing allergic inflammation under the continuous exposure of geohelminth infections. However, the loss of exposure due to helminth migration may reduce peripheral tolerance to active allergic responses. On the other hand, children born to uninfected mothers and live in areas with low geohelminth parasite transmission may have strong allergic responses to their first exposure to the parasites, and the effort to suppress the infection by anthelmintic therapy may paradoxically exacerbate allergic symptoms [6-8].

The frequency and patterns of helminth transmission from year to year are the major drivers of host immune responses to parasites, and they are linked to the natural immunological responses to helminth infections and allergies [3]. However, routine mass drug administration of anthelminthic in school-aged children has becoming a common practice to reduce the burden of STH infection in high prevalence areas, however the effect of treatment to the incidence allergic reactions are still contrasting [9]. In several districts in North Sumatera province, STH infections remain high from 55.8% in Sibolga City to 67.9% in Karo district [10-13]. This study aimed to determine the association between history of STH infections and incidence of positive skin prick test (SPT) in children receiving periodic anthelminthic treatment.

## Methods

This is a case-control study to evaluate the association between STH infections and allergic reactions based on SPT results among elementary school children. The study was done among school children aged 7 to 12 years old from January 2019 to February 2021. School children from Mandailing Natal district with recurrent STH infections (at least two positive results in 4-monthly Kato Katz examination over a year period) who received anthelminthic treatment following positive Kato Katz were classified as the case group, while school children aged 7 to 12 years old with no history of STH infections in the previous one year and a single negative Kato Katz examination from Medan were classified as the control group. Exclusion criteria were children who had recently or were experiencing allergic reactions (asthma attacks, urticaria, and anaphylaxis) or certain skin disorders (eczema or psoriasis), taking particular medications such as antihistamines, tricyclic antidepressants, beta blockers, and corticosteroids, and taking anthelmintic drugs (albendazole, mebendazole, pyrantel pamoate, and levamisole) in the past 4 weeks prior to each Kato Katz examination. Anthelminthic treatment was given according to species identification by Kato Katz examination, single dose of 400 mg albendazole for *A. lumbricoides* infection, and three days of 400 mg albendazole for single *T. trichiura* infection or mixed *A. lumbricoides* and *T. trichiura* infections.

Interviews and physical examinations were carried out according to the International Study of Asthma and Allergies in Childhood (ISAAC) questionnaire to all subjects to gather demographic data, nutritional status and history of previous medical condition, atopy, and family medical condition, as well as medication history. All subjects underwent skin prick test (SPT), where wheal size greater than or equal to 3 mm was considered positive. Allergens used were based on a previous study including histamine, negative control, *Dermatophagoides pteronyssinus, Blomia tropicalis*, American cockroach, cat dander, flower pollen (Mix-wildgrass), eggs (the entire egg), cow’s milk, and soy flour [14].

Data was analysed using a computerized software system SPSS version 22.0. Categorical data is presented as frequency and percentage distribution. Numerical data is shown in mean and standard deviation. The intervention in this research was the skin prick test (SPT). Chi-square, otherwise Fisher’s exact statistical analysis tests were performed to asses the role of STH infection in allergic reactions with 90% confidence interval (CI) and p<0.05 indicated significance.

The parents or legal guardians of all participants received participation information sheet, and written informed consent was obtained from their parents or legal guardians. This study was approved by the Health Research Ethical Committee, Universitas Sumatera Utara (Ref #57/TGL/KEPK FK USU-RSUPHAM/2020).

## Results

A total of 90 children were included in this study, each was 45 in the case and control groups. The mean age for children in the case and control groups were 9.4 (SD 1.3) and 8.7 (1.6), respectively. Eighty-nine percents of children in the case group were undernourished, while in contrast, only 15.6% were undernourished in the control group (Table 1). All children in the case group also reported of having a history of helminthiasis in the first interview, compared to none in the control group (100% vs 0%). On the other hand, none of the children in the case group had a history of allergy, while 40% of children in the control group had a history of allergy, with allergic rhinitis being the most common reported (*P*<0.001). History of parents and siblings with allergies, and their allergies diagnosis were also more significant in the control group (*P*<0.001, Table 2).

**Table 1.** Demographic characteristics.

| Demographic characteristics | Case<br>(n = 45) | Control<br>(n = 45) |
| --- | --- | --- |
| Gender, n (%) |  |  |
| Male | 18 (40) | 23 (51.1) |
| Female | 27 (60) | 22 (48.9) |
| Age, mean (SD), years old | 9.4 (1.3) | 8.7 (1.6) |
| Body weight, mean (SD), kg | 23.7 (6.4) | 28.9 (9.2) |
| Body height, mean (SD), cm | 129.9 (8.9) | 128.7 (10.7) |
| Nutritional status, n (%) |  |  |
| Undernourished | 40 (88.9) | 7 (15.6) |
| Normal | 5 (11.1) | 32 (71.1) |
| Overweight | 0 | 6 (13.3) |
| Socioeconomic status, n (%) |  |  |
| Poor | 39 (86.7) | 0 |
| Good | 6 (13.3) | 45 (100) |
| Regular anthelmintic drug consumption prior to the study, n (%) |  |  |
| Yes | 4 (8.9) | 45 (100) |
| History of helminthiasis, n (%) |  |  |
| Yes | 45 (100) | 0 (0) |
| Frequency of helminthiasis* |  |  |
| 2 times | 2 (4.4) | NA |
| 3 times | 31 (68.9) |  |
| 4 times | 12 (26.7) |  |

\*determined by Kato Katz examination, at 4 months interval
NA=not available

**Table 2.** The characteristics of allergies among study participants.

| Allergic characteristic | Case<br>(n = 45) | Control<br>(n = 45) | *p* |
| --- | --- | --- | --- |
| History of allergies, n (%) |  |  |  |
| No | 45 (100) | 18 (40) | <0.001<sup>a</sup> |
| Yes | 0 | 27 (60) |  |
| Type of allergies, n (%) |  |  |  |
| Asthma | 0 | 2 (4.4) | <0.001<sup>b</sup> |
| Allergic rhinitis | 0 | 22 (48.9) |  |
| Asthma and allergic rhinitis | 0 | 3 (6.70) |  |
| No disease | 45 (100) | 18 (40) |  |
| Parental allergies, n (%) |  |  |  |
| Yes | 0 | 26 (57.8) | <0.001<sup>a</sup> |
| No | 45 (100) | 19 (42.2) |  |
| Parents' medical condition, n (%) |  |  |  |
| Asthma | 0 | 8 (17.8) | <0.001<sup>b</sup> |
| Eczema | 0 | 4 (18.9) |  |
| Allergic rhinitis | 0 | 7 (15.6) |  |
| Asthma and eczema | 0 | 4 (8.9) |  |
| Asthma and allergic rhinitis | 0 | 3 (6.7) |  |
| No medical condition | 45 (100) | 19 (42.2) |  |
| Siblings with allergies, n (%) |  |  |  |
| Yes | 0 | 13 (28.9) | <0.001<sup>a</sup> |
| No | 45 (100) | 32 (71.1) |  |
| Siblings' medical condition, n (%) |  |  |  |
| Eczema | 0 | 3 (6.7) | 0.012<sup>b</sup> |
| Allergic rhinitis | 0 | 4 (8.9) |  |
| Eczema and allergic rhinitis | 0 | 1 (2.2) |  |
| Asthma and allergic rhinitis | 0 | 5 (11.1) |  |
| No medical condition | 45 | 32 (71.1) |  |

<sup>a</sup>Fischer's Exact, <sup>b</sup>Kruskal Wallis

There were 27% and 89% of children with positive skin prick test in the case and control groups, respectively, showing that positive skin prick test was more likely to occur among children in the control group (OR 22.0, 95% CI 7.0-68.8). Eight allergens were tested, and all was associated with increased positivity in children without history of STH infection (Table 3). Female was more likely to have positive SPT when exposed to cow’s milk allergen (OR 11.6, 95% CI 1.4-94.1), but no differences were seen in sex for other allergens. Other factors associated with increased risks of positive SPT were no history of helminthiasis, history of allergies, history of parents’ allergies, and history of siblings’ allergies (S1 Table). Allergic rhinitis increased the risk of positive SPT against *D. pteronyssinus* and *B. tropicalis*, but history of asthma did not increase the risk of positive SPT against any allergens tested.

**Table 3.** The association between positive SPT and STH infection.

| Skin prick test result | Case<br>(n = 45) | Control<br>(n = 45) | p* | OR<br>95%CI |
| --- | --- | --- | --- | --- |
| SPT, n (%) |  |  |  |  |
| Negative | 33 (73.3) | 5 (11.1) | <0.001<sup>a</sup> | 22.0 |
| Positive | 12 (26.7) | 40 (88.9) |  | 7.032-68.827 |
| Histamine, n (%) |  |  |  |  |
| Negative | 32 (71.1) | 10 (22.2) | <0.001<sup>a</sup> | 8.615 |
| Positive | 13 (28.9) | 35 (77.8) |  | 3.320-22.358 |
| *D. pteronyssinus*, n (%) |  |  |  |  |
| Negative | 44 (97.8) | 14 (31.1) | <0.001<sup>a</sup> | 97.429 |
| Positive | 1 (2.2) | 31 (68.9) |  | 12.170-779.99 |
| American cockroach, n (%) |  |  |  |  |
| Negative | 43 (95.6) | 21 (46.7) | <0.001<sup>a</sup> | 24.571 |
| Positive | 2 (4.4) | 24 (53.3) |  | 5.3-113.926 |
| Mix-wild grass, n (%) |  |  |  |  |
| Negative | 44 (97.8) | 29 (64.4) | <0.001<sup>a</sup> | 24.276 |
| Positive | 1 (2.2) | 16 (35.6) |  | 3.051-193.146 |
| Cow's milk, n (%) |  |  |  |  |
| Negative | 43 (95.6) | 35 (77.8) | 0.013<sup>a</sup> | 6.143 |
| Positive | 2 (4.4) | 10 (22.2) |  | 1.262-29.895 |
| *B. tropicalis*, n (%) |  |  |  |  |
| Negative | 43 (95.6) | 16 (35.6) | <0.001<sup>a</sup> | 38.969 |
| Positive | 2 (4.4) | 29 (64.4) |  | 8.324-182.424 |
| Cat dander, n (%) |  |  |  |  |
| Negative | 45 (100) | 25 (56.8) | <0.001<sup>b</sup> | - |
| Positive | 0 | 19 (43.2) |  |  |
| Eggs, n (%) |  |  |  |  |
| Negative | 45 (100) | 29 (64.4) | <0.001<sup>b</sup> | - |
| Positive | 0 | 16 (35.6) |  |  |
| Soy Flour, n (%) |  |  |  |  |
| Negative | 45 (100) | 39 (73.3) | <0.001<sup>b</sup> | - |
| Positive | 0 | 12 (26.7) |  |  |
| Negative control, n (%) |  |  |  |  |
| Negative | 45 (100) | 29 (64.4) | <0.001<sup>b</sup> | - |
| Positive | 0 | 16 (35.6) |  |  |

<sup>a</sup>Chi Square, <sup>b</sup>Fischer's Exact

## Discussions

This study compared the results of skin prick test in two areas with different prevalences of helminthiasis in children aged 7 to 12 years of age. Mandailing Natal has a high prevalence of STH infections among school-aged children, where STH prevalences were 100%, 78%, and 75% at 4-month intervals, and recurrent STH infections occurred in 65.4% of children despite periodic anthelminthic therapy. The socioeconomic status of the population was low, with only few households had personal sanitation available. Furthermore, children reported to have had poor hygienes by not having handwashing habit, inavailability of water and soap at schools, not wearing footwear, and self-reportedly play with dirt and soil daily (15,16). These factors have previously been reported to increase the incidence of STH infections [13,17-18].

In this study, children from Mandailing Natal district with at least two separate positive STH infections in the previous year, no history of allergy, nor parents’ and siblings’ allergies, had the SPT prevalence of 27%. In comparison, children from Medan City with no history of helminthiasis and negative Kato Katz examination, reported allergic symptoms (60%), and history of parents’ (57%) or siblings’ allergies (29%), showed a significantly higher of SPT prevalence (89%). The decreased exposure to common childhood infectious diseases and decreased diversity of microbial exposures in the environment, such as geohelminth (*A. lumbricoides, T. trichiura*, and hookworms) infections explains the rise in the prevalence of allergies [1,3]. Nevertheless, 27% of children with history of repeated STH infection and anthelminthic treatment in this study still revealed positive SPT results, although none reported any allergic symptoms. However, this proportion of SPT conversion of children who had received periodic anthelminthic is still much lower compared to previous studies from Indonesia (32%) and Gabon (69%) [19,20].

Chronic soil-transmitted helminth (STH) infections can modulate and suppress allergic reactions, particularly by changing the responses from the immune effector and by secreting immunomodulator cytokines that stimulate the protective dominant T-helper cell type-2 (Th2) to recruit the immune system cells such as eosinophils, basophils, mast cells, macrophages and differential production of immunoglobulin such as IgG1, IgG4, and especially IgE. These are similar responses as in allergic responses. Furthermore, the infection will activate anti-inflammatory responses including IL-10 and TGF-β where IL-10 has a potent antiallergic effect that can influence the response to allergen skin tests. IL-10 is essential in balancing the effectiveness of the immune system against persistent pathogenic infections and the illness the host has [5,6,21]. This study was done among children aged 7-9 years who might have had continuous exposure to STH during their early childhood, and a short-periodic treatment of anthelminthic might not be sufficient enough to reverse the protective effect of infections [22].

Food or protein allergies occur when the effective immune responses that are supposed to target parasites, target the protein instead, as mediated by the IgE. Antigen-presenting cells in the body pick up the protein or food, that has entered the body through ingestion, inhalation, and/or skin contact, triggering the allergies and offer them to the T-helper cells. Consequently, T-helper1 cells release cytokines and activate the cell-mediated responses, which are effective to fight against intracellular bacteria and protozoa. However, when the allergy-causing protein incorrectly stimulate T-helper2 cell responses, a type 1 hypersensitivity reaction may occur and the allergy responses by IgE may manifest in the skin, sinus, and/or lungs [5,6,14,23,24].

Previous study in 213 Southeast Asian children including Indonesian children has shown that the most common allergens identified were *Dermatophagoides pteronysinus* (70%), *Dermatophagoides farina* (69%), *Blomia tropicalis* (55%), dog hair (32%), cat dander (19%), and cockroach (19%). While the most common food allergies were to white eggs (54%), cow’s milk (31%), and soy flour (13%) [14]. As there was a limited study on which allergens causing allergies specific to Indonesian children, we used the results of their study to determine allergens used in our study. We found similar proportion of children with positive SPT to *D. pteronyssinus* and *B. tropicalis* although more children in our study were reacted to cockroach (53% vs 19%), cat dander (43% vs 19%), and mix wild grass (36% vs 7). Our study also showed higher proportion of allergies to eggs and cow’s milk than to soy flour. Different geographical, environmental, economical and cultural situation may affect the prevalence of allergies to certain allergens. The children in this study were from the upper-middle socioeconomic class who generally had good hygience level and were less likely to be exposed to pets like cats. Additionally, Medan as the area where the subjects in the control group were from is a densely populated area with a tropical climate and very few wild plants. Therefore, the initial exposure to wild plants, cockroach, and cat dander would stimulate rapid-type allergic reaction and activate Th-2 to produce IgE and trigger allergic reactions.

There were several limitations in our study. First, children in the control group had only a single examination for STH infection, therefore we could only confirm their absent history of helminthiasis based on the parents’ answers during the interview. However, our findings were supported by previous study in Medan among children from high socioeconomic status who had very low to no STH infection. Second, almost 90% of children with history of repeated helminthiasis were undernourished. Recurrent STH infections in children can lead to a number of bodily processes, including the penetration of gastrointestinal tract that impairs nutrient absorption and results in malnutrition. Malnutrition due to helminthiasis alters the regulation of IgE synthesis through the anti-inflammatory response activation, like IL-10 and TGF-β, in which IL-10 has a potent antiallergic effect that can modulate and suppress allergic reactions [5,6,25-28].

This study showed a contrast result of STH infection and SPT positivity among school-aged children in two areas in North Sumatera province. The findings are not only supported the hygiene hypothesis by revealing the association between infection exposures and allergies incidences in two contrasting areas, but also show no effect of anthelminthic treatment in reversing the protective effect of STH infections to allergies. We also show growing non-communicable diseases including allergies among children in big cities in developing countries, such as Indonesia. While programs to control and eliminate infectious diseases to support growth and development in children have been the main focuses for decades in many regions in Indonesia, transition from the communicable diseases-focused programs to the non-communicable disease prevention and management measures for children in big cities should also be commenced. Therefore, direct and long-term complications of non-communicable diseases in children can be prevented through lifestyle promotion and preventive measures.

## Conclusion

There were significant differences in characteristic between children without STH infections from Medan (case group) and children with STH infections from Mandailing Natal (control group). The research subjects without STH infections mostly had history of allergy compared to subjects with recurrent STH infections. There was a significant correlation between SPT and STH infections (p<0.001), where the group with recurrent STH infection mostly had negative SPT results.

## Data Availability

The data used to support the findings will be included in the article

## Acknowledgement

The authors would like to thank the participants for their participations, as well as the schools in Mandailing Natal and Medan city, especially the school principals and teachers for their support during the study.

## Supporting information

**S1 Table.** The relationship between subject’s characteristics and allergens based on skin prick test result

| Characteristic | Parasite | P | OR | 95% CI |
| --- | --- | --- | --- | --- |
| Gender. | *D. pteronyssinus* | 0.485<sup>a</sup> | 1.364 | 0.570-3.267 |
| Female | *A. cockroach* | 0.184<sup>a</sup> | 1.889 | 0.734-4.859 |
|  | *B. tropicalis* | 0.164<sup>a</sup> | 1.881 | 0.768-4.605 |
|  | Cow's milk | 0.005<sup>a</sup> | 11.579 | 1.425-94.057 |
| History of helminthiasis | *D. pteronyssinus* | <0.001<sup>a</sup> | 97.429 | 12.170-779.99 |
| Negative | *A. cockroach* | <0.001<sup>a</sup> | 24.571 | 5.300-113.926 |
|  | *B. tropicalis* | <0.001<sup>a</sup> | 38.969 | 8.324-182.424 |
|  | Cow's milk | 0.013<sup>a</sup> | 6.143 | 1.262-29.895 |
| History of allergy (+) | *D. pteronyssinus* | <0.001<sup>a</sup> | 23.320 | 7.144-76.121 |
|  | *A. cockroach* | <0.001<sup>a</sup> | 10.200 | 3.560-29.226 |
|  | *B. tropicalis* | <0.001<sup>a</sup> | 18.550 | 5.985-57.498 |
|  | Cow's milk | 0.021<sup>b</sup> | 4.060 | 1.157-14.244 |
| Asthma (+) | *D. pteronyssinus* | 0.054<sup>b</sup> | ~ |  |
|  | *A. cockroach* | 0.025<sup>b</sup> | ~ |  |
|  | *B. tropicalis* | 0.116<sup>b</sup> | ~ |  |
|  | Cow's milk | 0.016<sup>b</sup> | ~ |  |
| Allergic rhinitis (+) | *D. pteronyssinus* | <0.001<sup>a</sup> | 12.013 | 3.803-37.946 |
|  | *A. cockroach* | 0.006<sup>a</sup> | 4.629 | 1.661-12.895 |
|  | *B. tropicalis* | <0.001<sup>a</sup> | 9.422 | 3.138-28.292 |
|  | Cow's milk | 1.000<sup>b</sup> | 1.035 | 0.254-4.219 |
| Asthma and allergic rhinitis (+) | *D. pteronyssinus* | 0.042<sup>b</sup> | ~ |  |
|  | *A. cockroach* | 0.022<sup>b</sup> | ~ |  |
|  | *B. tropicalis* | 0.038<sup>b</sup> | ~ |  |
|  | Cow's milk | 0.046<sup>b</sup> | 15.400 | 1.278-185.599 |
| Parental allergies (+) | *D. pteronyssinus* | <0.001<sup>a</sup> | 14.444 | 4.773-43.715 |
|  | *A. cockroach* | <0.001<sup>a</sup> | 8.640 | 3.057-24.420 |
|  | *B. tropicalis* | <0.001<sup>a</sup> | 22.680 | 6.928-74.250 |
|  | Cow's milk | 0.004<sup>b</sup> | 6.667 | 1.798-24.726 |
| Siblings with allergies (+) | *D. pteronyssinus* | <0.001<sup>b</sup> | 34.200 | 4.177-280.018 |
|  | *A. cockroach* | 0.005<sup>b</sup> | 5.244 | 1.524-18.045 |
|  | *B. tropicalis* | <0.001<sup>b</sup> | 15.675 | 3.195-76.898 |
|  | Susu sapi | 0.046<sup>b</sup> | 3.833 | 0.958-15.340 |
| Socioeconomic status | *D. pteronyssinus* | <0.001<sup>b</sup> | ~ |  |
| Good | *A. cockroach* | <0.001<sup>a</sup> | 19.118 | 2.707-135.020 |
|  | *B. tropicalis* | <0.001<sup>a</sup> | 22.941 | 3.270-160.965 |
|  | Cow's milk | 0.001<sup>b</sup> | ~ |  |
| Latrine available | *D. pteronyssinus* | <0.001<sup>b</sup> | ~ |  |
|  | *A. cockroach* | <0.001<sup>a</sup> | 17.453 | 2.473-123.184 |
|  | *B. tropicalis* | <0.001<sup>a</sup> | 20.943 | 2.987-146.854 |
|  | Cow's milk | 0.001<sup>b</sup> | ~ |  |
| Does not play with soil | *D. pteronyssinus* | <0.001<sup>a</sup> | 31 | 4.419-217.451 |
|  | *A. cockroach* | <0.001<sup>a</sup> | 12 | 3.103-47.797 |
|  | *B. tropicalis* | <0.001<sup>a</sup> | 14.5 | 3.677-57.178 |
|  | Cow's milk | 0.013<sup>a</sup> | 5 | 1.160-21.549 |
| Clean nails | *D. pteronyssinus* | <0.001<sup>b</sup> | ~ |  |
|  | *A. cockroach* | <0.001<sup>a</sup> | 18.269 | 2.588-128.991 |
|  | *B. tropicalis* | <0.001<sup>a</sup> | 21.923 | 3.125-153.776 |
|  | Cow's milk | 0.001<sup>b</sup> | ~ |  |
| Good nutritional status | *D. pteronyssinus* | <0.001<sup>a</sup> | 4.297 | 2.175-8.492 |
|  | *A. cockroach* | <0.001<sup>a</sup> | 4.775 | 2.124-10.733 |
|  | *B. tropicalis* | <0.001<sup>a</sup> | 3.502 | 1.824-6.721 |
|  | Cow's milk | 0.220<sup>b</sup> | 2.005 | 0.689-5.835 |
| Consume anthelmintic | *D. pteronyssinus* | <0.001<sup>b</sup> | ~ |  |
|  | *A. cockroach* | <0.001<sup>a</sup> | 20.918 | 2.961-147.803 |
|  | *B. tropicalis* | <0.001<sup>a</sup> | 25.102 | 3.576-176.204 |
|  | Cow's milk | 0.005<sup>a</sup> | 9.204 | 1.240-68.326 |
| Wash hands | *D. pteronyssinus* | <0.001<sup>b</sup> | ~ |  |
|  | *A. cockroach* | <0.001<sup>a</sup> | 15.909 | 2.256-112.194 |
|  | *B. tropicalis* | <0.001<sup>a</sup> | 19.091 | 2.725-133.751 |
|  | Cow's milk | 0.025<sup>b</sup> | 7 | 0.945-51.867 |
| Wear footwear | *D. pteronyssinus* | <0.001<sup>b</sup> | ~ |  |
|  | *A. cockroach* | <0.001<sup>a</sup> | 14.474 | 2.055-101.961 |
|  | *B. tropicalis* | <0.001<sup>a</sup> | 17.368 | 2.482-121.552 |
|  | Cow's milk | 0.05<sup>b</sup> | 6.368 | 0.860-47.138 |
| History of helminthiasis I (-) | *D. pteronyssinus* | <0.001<sup>a</sup> | 22.654 | 3.233-158.734 |
|  | *A. cockroach* | <0.001<sup>a</sup> | 8.769 | 2.205-34.875 |
|  | *B. tropicalis* | <0.001<sup>a</sup> | 10.596 | 2.691-41.720 |
|  | Cow's milk | 0.054<sup>a</sup> | 3.654 | 0.849-15.724 |
| History of helminthiasis II (-) | *D. pteronyssinus* | <0.001<sup>a</sup> | 29.652 | 4.227-207.992 |
|  | *A. cockroach* | <0.001<sup>a</sup> | 11.478 | 2.882-45.717 |
|  | *B. tropicalis* | <0.001<sup>a</sup> | 13.870 | 3.517-54.691 |
|  | Cow's milk | 0.016<sup>a</sup> | 4.783 | 1.110-20.611 |
| History of helminthiasis III (-) | *D. pteronyssinus* | <0.001<sup>a</sup> | 10.029 | 1.452-69.266 |
|  | *A. cockroach* | 0.004<sup>a</sup> | 8.088 | 1.162-56.290 |
|  | *B. tropicalis* | <0.001<sup>a</sup> | 9.706 | 1.404-67.103 |
|  | Cow's milk | 0.280<sup>b</sup> | 3.559 | 0.487-26.031 |
| History of helminthiasis IV (-) | *D. pteronyssinus* | <0.001<sup>a</sup> | 24.800 | 3.537-173-865 |
|  | *A. cockroach* | 0.004<sup>a</sup> | 9.600 | 2.412-38.208 |
|  | *B. tropicalis* | <0.001<sup>a</sup> | 11.600 | 2.944-45.707 |
|  | Cow's milk | 0.038<sup>a</sup> | 4 | 0.929-17.226 |

<sup>a</sup>Chi Square. <sup>b</sup>Fischer's Exact

## References

1. Soegiarto G, Abdullah MS, Damayanti LA, Suseno A, Effendi C. The prevalence of allergic diseases in school children of metropolitan city in Indonesia shows a similar pattern to that of developed countries. Asia Pasific Allergy. 2019;9(2):1–10

2. Aranzamendi C, Sofronic ML, Pinelli E, Helminths: immunoregulation and inflammatory diseases which side are trichinella spp. and toxocara spp. on?. J Parasitol Res. 2013; 1–11

3. Xu F, Yan S, Li F, Cai M, Chai W, Wu M, et al. Prevalence of childhood atopic dermatitis: an urban and rural community-based study in Shanghai. China. PLoS One. 2012;7(5):e36174

4. Heinzerling L, Mari A, Bergman K, Bresciani M, Burbach G, Darsaw U, et al. The skin prick tes – European standards. Clin Transl Allergy. 2013;3(3):1–10

5. Redpath SA, Fonseca NM, Perona-Wright G. Protection and pathology during parasite infection: IL-10 strikes the balance. Parasite Immunol. 2014;36:233–52

6. Flohr C, Tuyen LN, Quinnell RJ, Lewis S, Minh TT, Campbell J, et al. Reduced helminth burden increases allergen skin sensitization but not clinical allergy: a randomized. double-blind. placebo-controlled trial in Vietnam. Clin Exp Allergy. 2010;40:131–142

7. Nampijja M, Webb EL, Kaweesa J, Kizindo R, Namutebi M, Nakazibwe E, et al. The Lake Victoria island intervention study on worms and allergy related diseases (LaVIISWA): study protocol for a randomised controlled trial. Bio Med Central. 2015;16(187):1–14

8. Cooper PJ. Mucosal immunology of geohelminth infection in humans. Mucosal Immunol. 2009;9(2):1–10

9. Sjafii RP, Pasaribu HS, Wijaya H, Pasaribu AP. Metode Pemberantasan kecacingan pada anak. J Indon Med Assoc. 2017;67(11):550–7

10. Pasaribu S. Penentuan optimal pengobatan massal askariasis dengan albendazole pada anak usia sekolah dasar di desa Suka. Pendekatan model dinamika populasi cacing. [Disertation]. [Medan]: Program Pasca Sarjana USU; 2004

11. Ginting A. Faktor-faktor yang berhubungan dengan kejadian kecacingan pada anak sekolah dasar di desa tertinggal kecamatan pangururan kabupaten samosir. [Thesis]. [Medan]: Fakultas Kesehatan Masyarakat USU; 2009

12. Pasaribu AP, Alam A, Sembiring K, Pasaribu S, Setiabudi D. Prevalence and risk factors of soil transmitted helminthiasis among school children living in an agricultural area of North Sumatera, Indonesia. BMC Public Health. 2019;19(1066):1–8

13. Pang KA, Pang KP, Pang EB, Cherilynn TYN, Chan YH, Siow JK. Food allergy and allergic rhinitis in 435 asian patients – A descriptive review. Med J Malaysia. 2017;72(4):215–20

14. Nasution RKA, Nasution BB, Lubis M, Lubis IND. Prevalence and knowledge of soiltransmitted helminth infections in Mandailing Natal, North Sumatera, Indonesia. Open Access Maced J Med Sci. 2019; 7(20):3443–6

15. Nasution RKA. Peranan edukasi berulang dengan media video film terhadap angka reinfeksi soil transmitted helminths pada anak Sekolah Dasar di dua kecamatan di Kabupaten Mandailing Natal, Provinsi Sumatera Utara. [Thesis]. [Medan]: Program Magister Kedokteran Klinik; 2021

16. Nery SV, Clarke NE, Richardson A, Traub R, McCarthy JS, Gray DJ, et al. Risk factors for infection with soil-transmitted helminths during an integrated community level water. sanitation. and hygiene and deworming intervention in Timor-Leste. Int J Parasitol. 2019;49(5):389–96

17. Bethony J, Brooker S, Albonico M, Geiger SM. Soil transmitted helminth infections: ascariasis, trichuriasis and hookworm. Lancet. 2006;367(9521):1521–32

18. Bogoch II, Andrews JR, Speich B, Ame SM, Ali SM, Stothard JR, et al. Short report: quantitative evaluation of a handheld light microscope for field diagnosis of soiltransmitted helminth infection. Am J Trop Med Hyg. 2014;91(6):1138–41

19. Wiria AE, Hamid F., Wammes LJ, Kaisar MMM, May L, Prasetyani MA, et al. The Effect of Three-Monthly Albendazole Treatment on Malarial Parasitemia and Allergy: A Household-Based Cluster-Randomized, Double-Blind, Placebo-Controlled Trial. PLoS One. 2013;8(3):1–9

20. van den Biggelaar AH, Rodrigues LC, van Ree R, et al. Long-term treatment of intestinal helminths increases mite skin-test reactivity in Gabonese schoolchildren. J Infect Dis. 2004;189(5):892-900. doi:10.1086/381767

21. Maizels RM, Hewitson JP, Smith KA. Susceptibility and immunity to helminth parasites. Curr Opin Immunol. 2012;24(4):459–66

22. Chico ME, Vaca MG, Rodriguez A, Cooper PJ. Soil-transmitted helminth parasites and allergy: Observations from Ecuador. Parasite Immunol. 2019;41(6):e12590. doi:10.1111/pim.12590

23. Alfredo FNJ, David RJ, Consuelo LM, Inés ML, Reyes P, Darío HR. Agreement of the kato-katz test established by the who with samples fixed with sodium acetate analyzed at 6 months to diagnose intestinal geohelminthes. Acta Trop. 2015;146:42–4

24. Hagel I, Lynch NR, Puccio F, Rodriguez O, Luzondo R, Rodríguez P, et al. Defective regulation of the protective ige response against intestinal helminth ascaris lumbricoides in malnourished children. J Trop Pediatr. 2003;49(3):136–42

25. McSorley HJ, Maizels RM. Helminth infections and host immune regulation. Clin Microbiol Rev. 2012;25(4):585–608

26. Ibrahim MK, Zambruni M, Melby CL, Melby PC. Impact of childhood malnutrition on host defense and infection. Clin Microbiol Rev. 2017;30(4):919–71

27. Rytter MJH, Kolte L, Briend A, Friis H, Christensen VB. The immune system in children with malnutrition—a systematic review. PLoS One. 2014;9(8):1–19

28. Wahyuni S, van Dorst MMAR, Amaruddin AI, Muhammad M, Yazdanbakhsh M, Hamid F, et al. The relationship between malnutrition and TH2 immune markers: a study in school-aged children of different socio-economic backgrounds in Makassar, Indonesia. Trop Med Int Health. 2021;26(2):195–203

